# Assessment of CEUS for evaluation of hepatic infantile hemangiomas

**DOI:** 10.1101/2025.05.08.25327084

**Authors:** G. Cericola, L. Gunst, H. Bel Hadj Jrad, J. Sahlmann, A. Puzik, A. Rahn, D. Franke, C. Voll, F.G. Kapp

## Abstract

**Background:** Infantile hemangiomas (IH) are the most common benign vascular tumor in infants. Despite their spontaneous regression, IH can cause aesthetic, functional, and vital organ complications. Hepatic involvement requires particular attention in infants with multiple cutaneous hemangiomas. While magnetic resonance imaging (MRI) is the established diagnostic standard, contrast-enhanced ultrasound (CEUS) emerges as a promising alternative.

**Objective:** To compare the diagnostic accuracy of CEUS and MRI in hepatic IH detection and characterization, and to evaluate potential diagnostic imaging markers.

**Materials and Methods:** A retrospective multicenter analysis was conducted to identify infants diagnosed with hepatic IH who underwent CEUS, MRI, or both between 2016 and 2024. Lesion size and count were compared in patients who underwent both imaging modalities. Data from 24 patients was included in the analysis.

**Results:** Of 24 patients, 15 underwent only CEUS, 3 underwent only MRI, and 6 received both imaging modalities. CEUS findings were comparable to MRI regarding lesion size and number, with CEUS demonstrating capability to detect lesions as small as 4 mm. Treatment decisions were predominantly based on CEUS findings.

**Conclusion:** CEUS showed diagnostic accuracy comparable to MRI in hepatic hemangioma evaluation, without compromising disease severity assessment. CEUS advantages include real-time imaging and avoidance of sedation or anesthesia in young patients. These findings support CEUS as a viable, less invasive, and resource-efficient alternative to MRI for hepatic IH evaluation, enabling optimization of diagnostic and treatment protocols.

**SUMMARY STATEMENT:** CEUS provides accurate diagnosis and monitoring of hepatic IH, demonstrating comparable or potentially superior detection capabilities to MRI, while offering practical advantages for infants.

**KEY RESULT:**

1. Diagnostic Accuracy: CEUS accurately detects lesion number and size.
2. Vascular Detailing: CEUS may offer superior real-time visualization of vascular patterns.
3. Clinical Benefits: CEUS is non-invasive, cost-effective, and suitable for repeated assessments in non-sedated patients, making it an ideal diagnostic modality for the assessment of suspected hepatic IH.

## INTRODUCTION

IH are benign vascular tumors affecting 4–5% of newborns, with higher prevalence in females and preterm infants. They typically emerge shortly after birth, undergo rapid proliferation during the first 4–6 months, followed by a plateau and gradual involution. By age 5, 50% have regressed, rising to 80% by age 8. Residual findings, including telangiectasias or fibrofatty tissue, may persist in some cases.^1^

The exact etiology remains unclear, but current theories suggest a multifactorial origin^2^. Recently, SOX18 and the mevalonate pathway have been implicated in IH development and as potential targets of R(+)-propranolol^3,4^.

Hepatic hemangiomas are classified into three distinct categories: focal, multifocal, and diffuse. Focal lesions, predominantly congenital rather than infantile, present as solitary, spherical tumors that are typically asymptomatic. In contrast, hepatic IH typically as multifocal or diffuse lesions and are associated with cutaneous hemangiomas. Multifocal lesions are composed of multiple well-defined nodules separated by normal liver tissue, while diffuse hemangiomas exhibit extensive hepatic replacement^5^. The diffuse form carries a higher risk of complications, including abdominal compartment syndrome and heart failure. These complications have become less common since the introduction of propranolol therapy^6–8^. While propranolol therapy has become the gold standard, diagnostic approaches remain non-standardized, resulting in significant practice variability, and potentially delaying diagnosis and treatment initiation.

Hepatic involvement should be suspected in infants with ≥5cutaneous hemangiomas^9^. It was shown that 77.4% of patients with multifocal and 53.3% with diffuse hepatic hemangiomas had concurrent cutaneous lesions^6^. In case hepatic lesions have been identified, magnetic resonance imaging (MRI) with and without contrast enhancement is often recommended but might lead to unnecessary exposure to gadolinium-based contrast agent in infancy. It further requires sedation or anesthesia and monitoring beds. CEUS using sulfur hexafluoride lipid-type A microspheres (Lumason/SonoVue®, Bracco Diagnostics) is a promising non-invasive alternative to assess hepatic lesions. Intravenously administered microbubbles, consisting of a gas core encapsulated by a stabilizing phospholipid shell, oscillate in response to ultrasound waves, enhancing acoustic scattering and producing strong echo signals. This mechanism enables high-resolution imaging and detailed visualization of the vasculature, including small vessels not accessible with conventional Doppler ultrasound. Lumason® (SonoVue®) is approved in Europe for various adult diagnostic applications, including liver imaging. While the U.S. FDA has approved its use for pediatric liver imaging, this indication remains off-label in Europe with its use increasing. Known contraindications include right-to-left cardiac shunts and severe pulmonary hypertension^10^.

CEUS presents a cost-effective alternative to MRI, potentially reducing diagnostic costs related to evaluation of hepatic IH^11–16^. However, to our knowledge, no larger cohort has been reported on its use to confirm the diagnosis of hepatic IH.

Here, we report on our multicentric experience with CEUS and MRI as imaging modalities for identifying hepatic IH. Through evaluating concordance between these imaging modalities, we seek to optimize the diagnostic approach for hepatic IH, potentially reducing reliance on more invasive or resource-intensive procedures and speeding up the diagnostic process while maintaining or even improving diagnostic accuracy.

## MATERIALS AND METHODS

### Patient Selection

This multicenter retrospective study reviewed CEUS and MRI examinations of patients with hepatic IH. Written informed consent was obtained from parents or legal guardians, and the study was approved by the institutional review boards of the University Medical Center Freiburg (21-1200) and Hannover (10527_BO_K_2022). Data collection spanned from October 1, 2016, to October 2024.

29 patients with suspected hepatic hemangiomas were initially identified. Five were excluded: three due to single lesions suggestive of congenital hemangioma, and two for incomplete clinical data. Thus, 24 patients met inclusion criteria (Figure 1).

**Figure 1:**
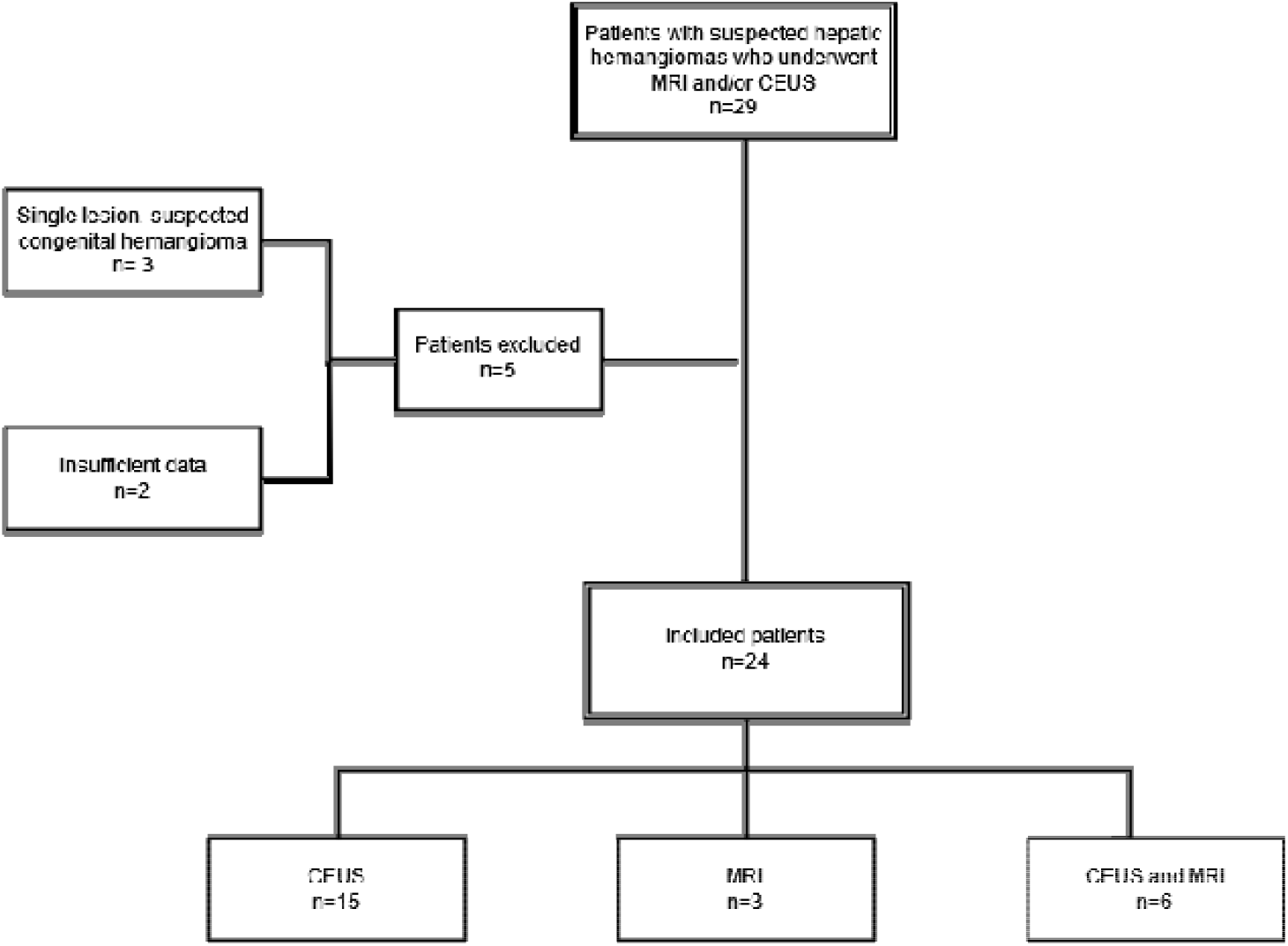
Study Popluation. This flow diagram depicts the patient selection process for our study on infantile hepatic hemangiomas

Collected data included lesion number, size, imaging features, severity of liver involvement, and Resistive Index (RI). Clinical profiles comprised age, sex, prematurity, and laboratory results (tumor markers, thyroid hormones, proBNP). Treatment and outcomes were also recorded.

### Contrast-enhanced ultrasound (CEUS)

CEUS exams were performed using different systems: Zonare ZS3 Mindray in Freiburg; GE Logiq E10, E9, S8, and Toshiba Aplio XG in Hannover. SonoVue® contrast agent was administered based on pediatric dosing guidelines: 0.1 mL/year of life or 0.03 mL/kg, followed by a 10 mL saline flush. Imaging included a 20-second dynamic sequence post-injection and static captures at one-minute intervals for 5–6 minutes. IH diagnosis was based on typical enhancement patterns: early peripheral nodular enhancement with centripetal fill-in during the arterial phase, progressing to homogeneous enhancement in later phases. RI was measured by pulsed-wave Doppler within the main hepatic artery using the formula: (Peak Systolic Velocity - End Diastolic Velocity) / Peak Systolic Velocity, averaged over three waveforms. An experienced pediatric sonographer interpreted all CEUS exams.

### Magnetic resonance imaging examination

Four patients underwent contrast-enhanced MRI using T1-weighted Turbo Spin Echo (TSE) and STIR sequences with ProHance® (Gadoteridol). Hemangiomas showed typical peripheral nodular enhancement with delayed central filling, hyperintensity on T2-weighted images, and no signal on pre-contrast T1 sequences. No diffusion restriction was observed, supporting benignity. The remaining five patients underwent non-contrast MRI on 1.5T or 3.0T scanners. Protocols included axial and coronal T1-weighted sequences, fat-suppressed T2-weighted imaging, and diffusion-weighted imaging with b-values of 50, 400, and 800 s/mm^2^, alongside ADC map calculation. In-phase and out-of-phase sequences were used for fat content analysis. An experienced pediatric radiologist evaluated all MRI images, documenting abnormal enhancement and notable findings in T1 and STIR sequences.

### Statistical Analysis

CEUS and MRI lesion characterization agreement was assessed using scatter plots and Spearman’s rank correlation coefficient (ρ), comparing lesion size by CEUS (x-axis) and MRI (y-axis). Spearman’s ρ was chosen for its suitability in analyzing ordinal data and monotonic relationships, with values ranging from −1 (perfect negative correlation) to +1 (perfect positive correlation). To examine the relationship between RI and lesion characteristics, a multiple linear regression was conducted with RI as the dependent variable, and lesion number and size as independent variables. Analysis was performed using Microsoft Excel’s regression tool, with significance set at p < 0.05.

Given the limited sample size (n=11), results should be interpreted cautiously. Multicollinearity was not formally assessed, and regression assumptions (linearity, independence, homoscedasticity, normality of residuals) were not rigorously tested. Thus, the findings are preliminary and warrant confirmation in larger cohorts.

## RESULTS

### Patient Characteristics

Patient characteristic depicted in tables 1 and 2. This multicenter study analyzed 24 infants diagnosed with hepatic IH. 23 patients had multifocal hepatic hemangiomas, while one (P20) presented with diffuse involvement. There was a female predominance with 18 females (75%) Age at diagnosis ranged from birth to eight months (median: 3 months) (Tables 1 and 2).

**Table 1.** Patient characteristics. This table summarizes the demographic and clinical data for the patients involved in the study. Abbreviations are as follows: mo: month, F: female, M: male, Y: yes, N: no, n/a: not available. The columns provide the age of the patient at the time of diagnosis, sex, prematurity and gestational age, and whether cutaneous hemangiomas are present. Additional relevant details, such as the presence of other conditions, are also included.

|  | Sex | Multifocal cutaneous IH |
| --- | --- | --- |
| P1 | F | Y |
| P2 | F | Y |
| P3 | F | Y |
| P4 | M | N |
| P5 | F | Y |
| P6 | F | Y |
| P7 | F | Y |
| P8 | F | Y |
| P9 | F | N |
| P10 | F | Y |
| P11 | F | Y |
| P12 | F | Y |
| P13 | M | n/a |
| P14 | F | Y |
| P15 | F | Y |
| P16 | M | Y |
| P17 | F | Y |
| P18 | F | n/a |
| P19 | F | n/a |
| P20 | F | n/a |
| P21 | F | Y |
| P22 | M | n/a |
| P23 | M | Y |
| P24 | M | n/a |

**Table 2.**
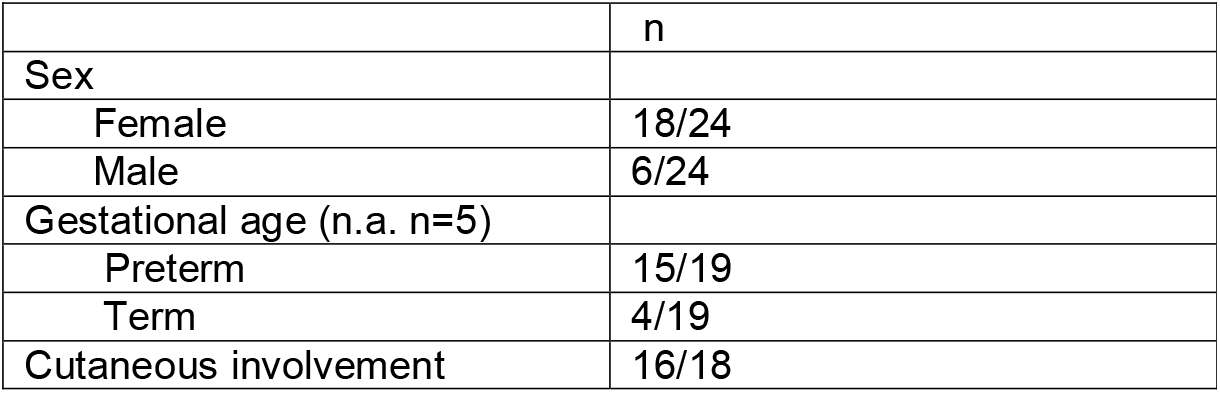
The table summarizes the demographic and clinical characteristics of the study population, including sex distribution, gestational age, and cutaneous involvement, along with their respective statistical significance (p-values).

|  | n |
| --- | --- |
| Sex |  |
| Female | 18/24 |
| Male | 6/24 |
| Gestational age (n.a. n=5) |  |
| Preterm | 15/19 |
| Term | 4/19 |
| Cutaneous involvement | 16/18 |

Nine patients (37.5%) were diagnosed between birth and two months, five (20.8%) between three and four months, nine (37.5%) between five and six months, and one (4.1%) between seven and eight months.

Gestational age was available for 19 patients: 15 were preterm (78.9%) and four at term (21%). Prematurity ranged from 25 ^4^/_7_ weeks (P12) to 36 ^5^/_7_ weeks (P11).

Regarding cutaneous involvement, data was available for 18 patients. Among these, 16 patients (88.9%) had multifocal cutaneous IH (p = 0.001), while P4 and P9 had no skin involvement (11.1%). P11 had a central nervous system IH.

### Laboratory Findings

All laboratory finding are depicted in table 3. AFP was within age-adjusted reference values in all tested patients (n=14) ^17^.

**Table 3.**
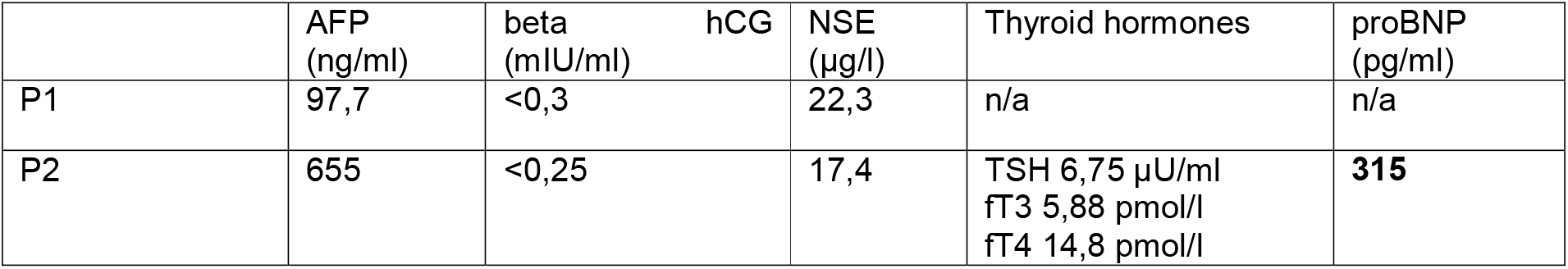
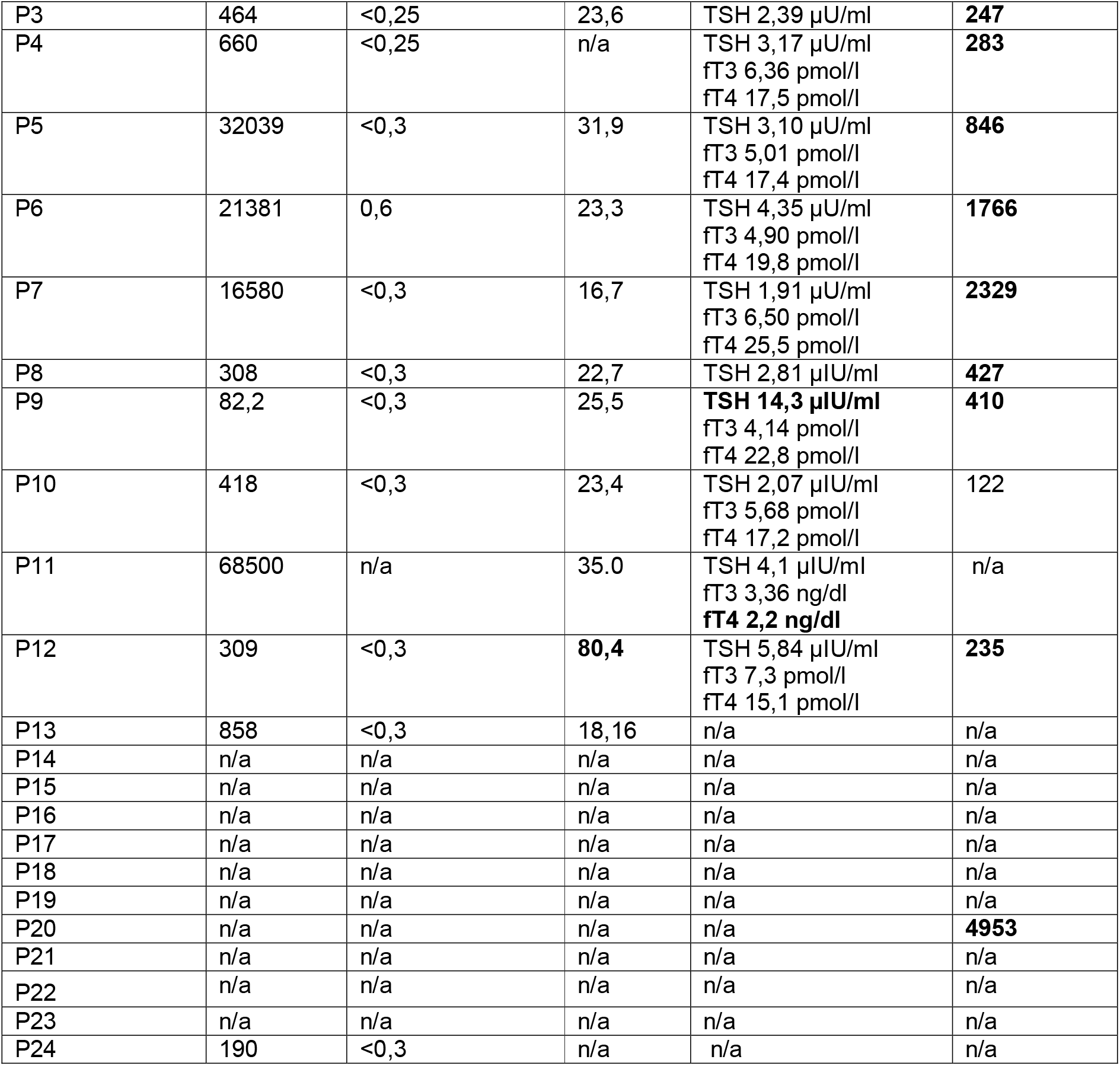
Lab Results at Diagnosis. This table presents the lab results for each patient at the time of diagnosis. The reference values used for comparison are as follows: TSH 0.73-8.35 µU/ml, fT3 3.30 −8.95 pmol/l, and fT4 11.9-25.6 pmol/l. NSE <17 µg/l and Beta-HCG <5.3 mIU/ml. In neonates, reference values for proBNP are not well established. The reference value for AFP was based on Blohm et al^17^.

NSE was assessed in 12 patients. Two had normal values (<17 µg/L). Mild elevation (17.4– 35 µg/L) was observed in 10 patients. P12 had a pathological level (80.4 µg/L), but urine catecholamines were normal.

Beta-hCG was measured in 13 patients, all within the normal range (<5.3 mIU/ml).

Thyroid parameters were analyzed in 11 patients, with nine showing unremarkable levels. One patient presented with elevated TSH level above the reference range (0.73-8.35 µU/ml). P9, with extensive liver involvement (>10 lesions, 40-45 mm in diameter), showed markedly elevated TSH (36.8 µU/ml), with normal free T3 (4.14 pmol/L), and T4 (22.8 pmol/L). P11, again with extensive liver involvement (>10 lesions, 12.8 mm in diameter), demonstrated slightly reduced free T4 levels at 2.2 ng/dL.

ProBNP levels were measured in 11 patients. It is important to note that in neonates, reference values for proBNP are not well established. Levels are typically elevated shortly after birth and decline physiologically within the first 1–3 months of life. Only P10 had a value <125 pg/ml. Others ranged from 235 pg/ml (P12) to 4953 pg/ml (P20) (median 410 pg/mL. P6 and P7 also had elevated values (1766 and 2329 pg/ml) but normal echocardiography. P20, with diffuse IH and the highest proBNP, had an enlarged atrium and ventricle, impaired systolic function, and elevated right ventricular pressure. After propranolol, cardiac findings normalized.

### Imaging Findings

All imaging findings are summarized in table 4. This retrospective analysis included data from 24 patients who underwent imaging for liver lesion characterization. CEUS was performed in 21 patients, MRI in nine patients. Both imaging modalities were performed in six patients, enabling direct comparison between modalities. Among the 21 patients examined with CEUS, maximum lesion diameters ranged from 4 to 45 mm. CEUS detected fewer than five lesions in one patient, five to ten lesions in four patients, and more than 10 lesions in 15 patients. In the nine patients who underwent MRI, three patients had 5-10 lesions, while six patients had more than 10 lesions. The six patients who underwent both imaging modalities demonstrated 100% concordance in lesion number. Due to perfect agreement, Cohen’s Kappa could not be calculated (due to division by zero), but a statistical significant concordance can be assumed. Lesion size measurements revealed only minor variations between modalities in three patients: P3 and P10 had only slight variations (13 mm CEUS vs. 10 mm MRI, and 11 mm vs. 15 mm, respectively), while P5 exhibited the most notable difference (10 mm vs. 3–5 mm), with CEUS suggesting a larger lesion size. The Spearman’s rank correlation coefficient (ρ) of 0.943 indicated a very strong positive correlation between the two imaging modalities, supported by the scatter plot (Figure 2). Hepatic artery RI was measured in 11 patients with a median of 0.69 (range 0.59–0.80), with one patient showing mild elevation (P20, with diffuse hepatic hemangioma), and one patient with pathological elevation (P15). No statistically significant correlation was observed between RI values and liver involvement (ρ_size_ = −0.1696, ρ_number_ = 0.0180), indicating that neither lesion size nor lesion number have a meaningful monotonic relationship with RI.

**Table 4.** Lesion Characteristics in CEUS and MRI. This table provides the maximum diameter and number of lesions observed in CEUS(CEUS) and magnetic resonance imaging (MRI) for each patient. Additionally, the RI (RI) of the hepatic artery is listed, normal range 0.5 – 0.75 pathologic RI <0.5 und >0.8.^35^

|  | max. diameter<br>CEUS (mm) | max. diameter<br>MRI (mm) | number of<br>lesions CEUS | number of<br>lesions MRI | RI A.hep. |
| --- | --- | --- | --- | --- | --- |
| P1 | 11-12 | n/a | 5-10 | n/a |  |
| P2 | 8-9 | n/a | >10 | n/a |  |
| P3 | 13 | 10 | >10 | >10 |  |
| P4 | 6-7 | 6 | >10 | >10 |  |
| P5 | 10 | 3-5 | 5-10 | 5-10 |  |
| P6 | 7 | 7 | >10 | >10 |  |
| P7 | n/a | 8 | n/a | 5-10 |  |
| P8 | 5 | n/a | >10 | n/a |  |
| P9 | 40-45 | 40-45 | >10 | >10 | 0.67 |
| P10 | n/a | 9 | n/a | 5-10 |  |
| P11 | n/a | 12,8 | n/a | >10 |  |
| P12 | 4 | n/a | <5 | n/a |  |
| P13 | 11 | 15 | >10 | >10 | - |
| P14 | 8 | n/a | >10 | n/a | 0.69 |
| P15 | 6 | n/a | >10 | n/a | 0.8 |
| P16 | 20 | n/a | >10 | n/a | 0.65 |
| P17 | 9 | n/a | 5-10 | n/a | - |
| P18 | 16 | n/a | 5-10 | n/a | 0.68 |
| P19 | 10 | n/a | >10 | n/a | 0.69 |
| P20 | 30 | n/a | >10 | n/a | 0.78 |
| P21 | 14 | n/a | >10 | n/a | 0.74 |
| P22 | 8 | n/a | >10 | n/a | 0.59 |
| P23 | 18 | n/a | >10 | n/a | 0.72 |
| P24 | 19 | n/a | >10 | n/a | 0.72 |

**Figure 2:**
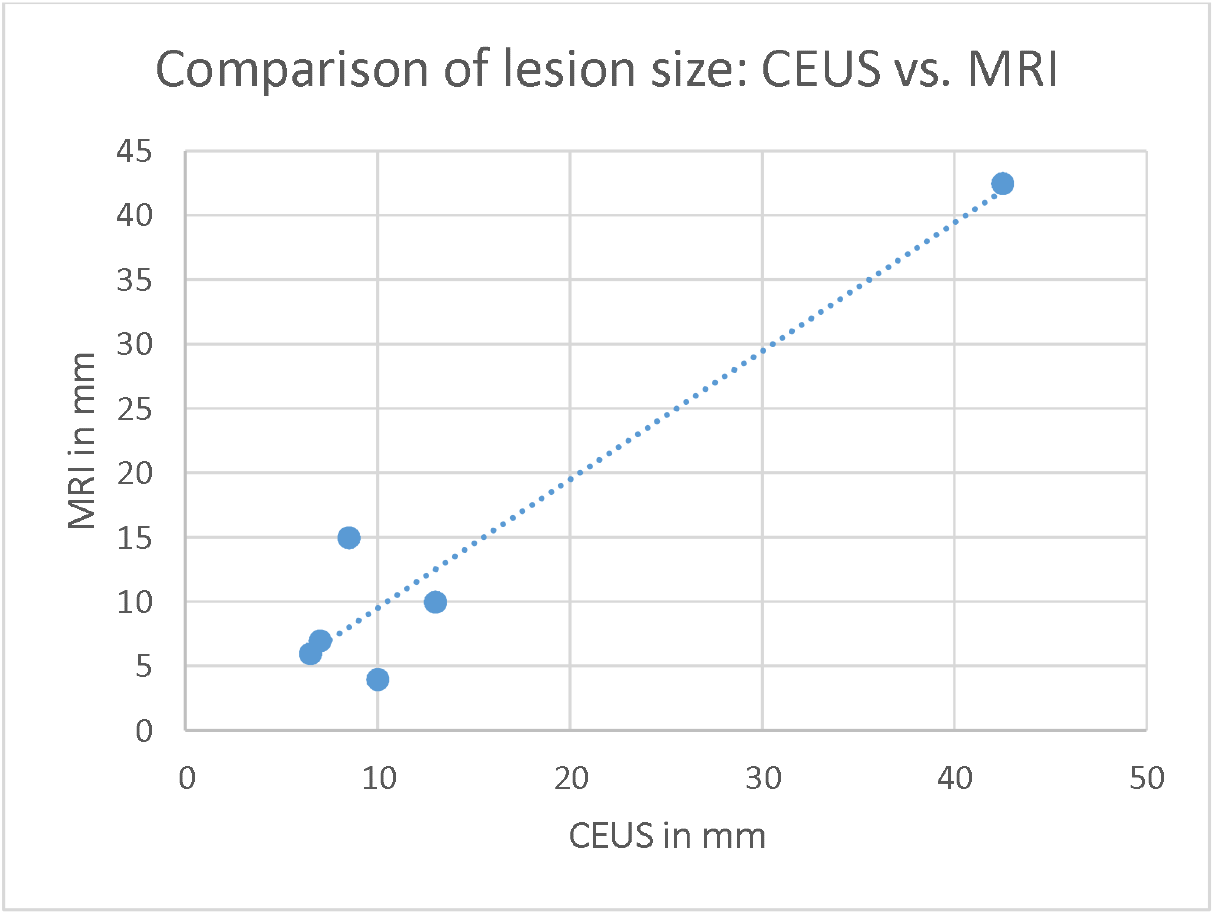
Comparison of lesion measurements obtained via CEUS and MRI. Scatter plot illustrating the correlation between lesion sizes (in mm) as measured by CEUS (x-axis) and MRI (y-axis). A dotted line represents a trend of increasing lesion size in MRI with increasing lesion size in CEUS.

### Treatment Outcomes

Treatment outcomes are depicted in table 5. 19 patients were evaluated for therapy duration and treatment response. Propranolol was the primary treatment, administered to 17 patients. Six patients were treated for five to 14 months, five were still under therapy at analysis, and six lacked complete data. Complete response to treatment was documented in nine patients. four patients showed significant regression during treatment with residual lesion sizes of 3 and 4 mm (not documented in two patient). Two patients achieved complete regression under an observational approach. P21 showed regression on CEUS, but treatment details were unavailable.

**Table 5.** Therapy and Remission Status. This table summarizes the treatment applied, duration of therapy, and the outcome for each patient. The outcome indicates whether a complete response or regression occurred.

|  | Therapy, duration | Outcome |
| --- | --- | --- |
| P1 | Propanolol, 5 mo | Regression (3 mm) |
| P2 | Propanolol, 10 mo | Complete response |
| P3 | Propanolol, 7 mo | Regression (4 mm) |
| P4 | No therapy | Complete response |
| P5 | Propanolol, 9 mo | Complete response |
| P6 | Propanolol, duration unknown | Complete response |
| P7 | Propanolol, 12 mo | Complete response |
| P8 | Propanolol, ongoing | n/a |
| P9 | Propanolol, ongoing | n/a |
| P10 | Propanolol, ongoing | n/a |
| P11 | Propanolol, ongoing | Regression |
| P12 | Propanolol, ongoing | n/a |
| P13 | Propanolol suggested | n/a |
| P14 | Propanolol, duration unknown | Regression |
| P15 | No therapy | Complete response |
| P16 | n/a | n/a |
| P17 | n/a | Complete response |
| P18 | Propanolol, duration unknown | Complete response |
| P19 | n/a | n/a |
| P20 | Propanolol, 14 mo | Complete response |
| P21 | n/a | Regression |
| P22 | Propanolol, duration unknown | Complete response |
| P23 | Propanolol, duration unknown | Complete response |
| P24 | Propanolol, duration unknown | n/a |

## DISCUSSION

Hepatic involvement is a rare manifestation of multifocal IH. In this study, we evaluated patient characteristics, laboratory parameters and imaging modalities in patients with hepatic IH to provide the basis for a non-invasive, fast and accurate diagnosis using CEUS.

A striking finding in our cohort was the robust correlation between hepatic IH and prematurity. This observation aligns with previous research^5^ and emphasizes the importance of screening in infants with multiple cutaneous hemangiomas, especially with a history of prematurity.

Cardiac function was impaired in only one patient in our cohort, evident by excessively elevated proBNP levels and echocardiographic findings. Cardiac function normalized during treatment. ProBNP serves as a highly sensitive marker of cardiac volume overload, detecting changes before echocardiographic abnormalities become apparent. However, reference values are lacking in neonates and elevation is common in the first 1-3 months of life due to increased pulmonary resistance. ProBNP elevation may also be caused by diffuse hepatic hemangiomas causing high-output cardiac failure^18,19^. Our observations suggest that in cases of extensive liver involvement, proBNP determination and subsequent echocardiography in case of excessively elevated values or clinical signs of cardiac insufficiency is warranted.

Thyroid dysfunction may occur in IH with extensive hepatic disease due to type 3 iodothyronine deiodinase activity in hemangioma tissue, inactivating thyroid hormones^20,21^. Newborn screening may fail to detect this complication, as it typically develops after hemangioma proliferation in the early weeks or months of life. Our findings thus support measuring thyroid hormone levelsin patients with hepatic IH^22^.

Assessment of tumor markers was unremarkable in our highly selected patient cohort. However, these remain valuable in patients with more ambiguous findings and relevant differential diagnoses of liver lesions^23,24^.

To explore predictive imaging parameters, we analyzed hepatic artery resistive index (RI). While values were typically in the high-normal range, only two exceeded the upper limit and RI did not correlate with lesion size or number, limiting its predictive value for therapeutic intervention. Only one patient in our cohort had a constellation of findings suggesting altered hemodynamics, including elevated proBNP, volume overload on echocardiography, and elevated hepatic artery RI. In future studies, peak systolic velocity (PSV) of the hepatic artery and hepatic vein dilation may be assessed to stratify patients with a need for therapy more objectively. These parameters were identified as significant predictors of complications in a previous study involving 112 patients with various liver lesions, including focal, multifocal, and diffuse hepatic hemangiomas, as well as malignant tumors^25^.

Our experience with CEUS confirmed its reliability. While CEUS visualization is limited to a specific liver window during examination, combining it with conventional ultrasound enables comprehensive assessment of both lesion number and diameter. Concordance with MRI in lesion number was 100%, and lesion size showed strong agreement and in the only case of differing size measurements, CEUS estimated a larger lesion size than MRI. Our observation aligns with studies reporting superior detection of arterial hypervascularity of CEUS in adult liver lesions, due to real-time assessment^26^. The enhanced sensitivity to vascular structures suggests that CEUS may provide more precise measurements of highly vascularized lesions like hemangiomas, potentially surpassing MRI. Our findings suggest that CEUS does not underestimate the extent or severity of hepatic lesions compared to MRI.

Recent studies have demonstrated CEUS’ effectiveness in characterizing and stratifying focal liver lesions in children. These focused on various lesions, including hemangiomas, focal nodular hyperplasia (FNH), hepatocellular adenomas, and malignant tumors such as hepatoblastomas and metastases. Key findings indicate that early washout (≤45 seconds) strongly suggests malignancy, particularly for hepatoblastoma in patients under five years old. Key findings indicate that early washout (≤45 seconds) strongly suggests malignancy, particularly hepatoblastoma in children under five. Hemangiomas are characterized by peripheral discontinuous globular hyperenhancement with centripetal fill-in. CEUS demonstrated high sensitivity (84.6–90.7%) and specificity (93.6–100%) in differentiating benign from malignant lesions^10,27–29^.

In addition to diagnostic accuracy, CEUS offers significant advantages in pediatric liver imaging as it is safe, efficient, does not require sedation, allows parental presence during the exam, and is readily available^30–32^. The contrast agent is eliminated via the lungs, does not accumulate in the body and has no nephrotoxic effects^32^. We believe that in an infant with multifocal cutaneous IH and multiple liver lesions, CEUS is the optimal imaging modality to make the diagnosis of hepatic IH. MRI should be reserved for patients with ambiguous findings to avoid gadolinium exposure at this young age, due to recent studies showing gadolinium accumulation in brain and other tissues, raising concerns about potential long-term toxicity, especially in the developing brains of children^33^. Additionally, GBCA have been associated with nephrogenic systemic fibrosis in patients including children with impaired renal function, although this risk has been significantly reduced with current guidelines^33,34^.

Because venous access is needed for CEUS, we currently recommend to check thyroid function parameters (especially patients with extensive hepatic lesions). Measurement of additional laboratory parameters depends on the extent of hepatic lesions and likelihood of differential diagnoses and may include proBNP (e.g. in patients with clinical signs of cardiac insufficiency or with extensive liver lesions) and tumor markers (e.g. in patients with a lack of cutaneous hemangiomas or atypical ultrasound and CEUS findings).

Follow-up is recommended for both treated and untreated patients to confirm hepatic lesion regression. In our experience, hepatic IH regress more slowly under therapy than cutaneous IH.

This study has several limitations. Its retrospective nature and its relatively small sample size restricts generalization of its results. Additionally, CEUS is dependent on an experienced examiner. Due to the rarity of this disease and the existence of relevant differential diagnoses, patients with suspicion of hepatic IH should be referred to a specialized center.

In conclusion, CEUS combined with conventional ultrasound demonstrates major advantages in hepatic IH imaging, providing comprehensive and accurate assessment with consistent detection of lesion number and size. Its enhanced sensitivity to vascular structures may enable more precise evaluation than MRI while offering a safe, fast, and efficient diagnostic tool. Our findings support CEUS as a first-line modality in evaluating suspected hepatic IH, especially where MRI access is limited, sedation is concerning, or gadolinium should be avoided. Future research should focus on prospective studies to validate these findings and establish standardized CEUS protocols.

## Data Availability

All data produced in the present study are available upon reasonable request to the authors

## Acknowledgements

The authors first and foremost acknowledge the patients and their families for providing their information and allowing us to conduct this work. We further acknowledge the Center for Vascular Anomalies at the Freiburg Center for Rare Diseases. Five of the authors are members of the Vascular Anomalies Working Group (VASCA WG) of the European Reference Network for Rare Multisystemic Vascular Diseases (VASCERN), and of the German Reference Network for Vascular Anomalies. FK was supported by the charity cycling tour “Tour der Hoffnung” and received funding from the German Federal Ministry of Education and Research (BMBF): EJPRD Joint Transnational Call 2023, NARRATIVE;FKZ 01GM2405).

## Conflicts of interest

Friedrich G. Kapp has received consulting fees from Novartis.

## Notes

### Funding Statement

This study did not receive any funding

### Author Declarations

Ethics committee of University Medical Center Freiburg (21-1200) and Hannover (10527_BO_K_2022) gave ethical approval for this work

